# The longitudinal care cascade for hypertension: a clinic-based study of people with and without HIV in South Africa

**DOI:** 10.64898/2026.02.11.26346061

**Authors:** S. Gumede, J. Manne-Goehler, E. Oladimeji, N. Bulled, A.T Brennan, S. Lalla-Edward

## Abstract

**Background:** Hypertension (HTN) constitutes a major and growing public health challenge in South Africa, a setting where HIV prevalence also remains high. Despite this dual burden, longitudinal evidence describing how individuals move through the HTN care cascade particularly comparing people living with HIV (PLHIV) and people not living with HIV (PNLHIV) remains limited. Understanding patterns of progression and regression across the cascade is essential to inform health system strategies for integrated chronic disease management.

**Methods and Findings:** We conducted a longitudinal secondary analysis using data collected from the iHEART-SA trial, which was implemented across nine public primary healthcare clinics in Johannesburg between August 2022 and May 2024. with follow-up through May 2025. Adults (≥18 years) with a known HIV status and a completed medical file review were included. Progression and regression were assessed across the HTN care cascade.

Of 23 855 participants, 78.5% were PLHIV (median age 42 years, IQR 36–49; 69% female). Overall, 34.4% had high blood pressure (BP), with prevalence higher among PNLHIV than PLHIV (53.5% vs 29.3%; p<0.001). Along the HTN care cascade, PLHIV were substantially more likely to remain undiagnosed compared with PNLHIV (aRR 3.40; 95% CI 3.12–3.71). Among those treated, PLHIV were less likely to achieve BP control (aRR 0.83; 95% CI 0.75–0.91). During follow-up, PLHIV experienced higher rates of cascade regression, including regression to untreated HTN (29% vs 19%; aRR 1.51; 95% CI 1.35–1.70) and from controlled to uncontrolled BP (aRR 1.12; 95% CI 1.05–1.18).

**Conclusions:** Despite frequent health-system contact, PLHIV had lower HTN diagnosis and higher regression, including treatment discontinuation and loss of BP control, underscoring persistent gaps in longitudinal HTN management within HIV programmes and the need for integrated and targeted chronic care models that ensure treatment continuity and sustained control.

## Background

Hypertension (HTN) is a growing public health challenge in middle-income countries (MICs) [1–3]. In South Africa, the prevalence of HTN is approximately 46% in the general population, with estimates suggesting similar or slightly lower rates among people living with HIV (PLHIV) compared to people not living with HIV (PNLHIV) [3,3–6]. The convergence of the HIV and HTN epidemics presents a significant challenge for the health system but also offers an opportunity to integrate chronic disease management. Several cross-sectional studies have suggesFted that HTN care may be more effective among PLHIV, largely due to more frequent clinical contact and better continuity of care through established HIV programmes [2,3,7–9]. The robust HIV care infrastructure facilitates systematic screening, earlier treatment initiation, and regular follow-up for comorbid conditions such as HTN. Consequently, PLHIV may have higher rates of diagnosis and treatment initiation for HTN compared to PNLHIV. However, this differential access could also widen disparities in HTN care outcomes between these populations if similar systems are not available for PNLHIV.

Despite the feasibility of treating and controlling HTN within the South African healthcare system, current evidence suggests that effective blood pressure (BP) control remains suboptimal at the population level [10,11]. Cross-sectional data indicate that only about 10% of HTN individuals achieve control (often summarized as the “ABC” — Awareness, BP control, and Compliance) [11]. Sustaining this control over time is even more challenging; a longitudinal study from South Africa found that only around 51% of patients who initially achieved control maintained it at follow-up [12]. However, there is limited evidence on whether such patterns differ by HIV status, particularly over time.

To date, longitudinal data directly comparing progression through the HTN care cascade, including diagnosis, treatment, and sustained BP control between PLHIV and PNLHIV are lacking [11,13,14]. Generating such evidence is critical for informing health system interventions that leverage the strengths of existing HIV care platforms to improve cardiovascular outcomes more broadly. These insights are especially relevant in South Africa, where integrated chronic disease care models are being scaled up to address the dual burden of infectious and non-communicable diseases (NCDs).

In this study, we use a large clinical dataset from the Integrating HIV and Heart Health in South Africa (iHEART-SA) trial to provide a comprehensive description and comparison of progress through various stages of the HTN care cascade among PLHIV and PNLHIV in Johannesburg, South Africa.

## Methods

### Study setting

The iHEART-SA trial was conducted in 9 of 14 public sector clinics in region F (inner-city) of the City of Johannesburg, a district within the Gauteng province. These clinics serve almost 700,000 residents in total and offer reproductive and sexual health care, mobile and community outreach, HIV screening and treatment, and tuberculosis screening and treatment. As the economic centre of the country, Johannesburg has a large immigrant population, including both South Africans from different provinces within the country and individuals from other countries.

### Data source

This longitudinal study was conducted as a secondary analysis of data from the iHEART-SA trial. The iHEART-SA project (Human Research Ethics Committee [HREC] number: M211160) was designed to evaluate an intervention to improve the diagnosis and management of HTN and to strengthen delivery of recommended HTN care within selected public primary healthcare clinics in Johannesburg, South Africa. The trial employed a 24-month cluster stepped-wedge implementation-effectiveness type 2 hybrid design across nine clinics, simultaneously assessing implementation outcomes at the provider and clinic level and effectiveness outcomes at the patient level [15].

The iHEART-SA trial defined two analytic populations. The implementation evaluation population included all adults (≥18 years) presenting for care at participating clinics who were seen in the triage area where vital signs were routinely measured, irrespective of HIV status or BP. These individuals contributed data to implementation outcomes across both control and intervention periods through routine data collection at triage.

Within this clinic-based population, the effectiveness evaluation population comprised adults aged ≥18 years with a known diagnosis of HIV who were observed to have elevated BP (systolic ≥140 mmHg or diastolic ≥90 mmHg) at any point during the control and intervention periods and who consented to prospective medical record abstraction. The primary iHEART-SA effectiveness analyses were therefore restricted to PLHIV, consistent with the trial’s eligibility criteria.

Although the parent trial’s effectiveness analyses focused exclusively on PLHIV, PNLHIV who attended participating clinics and consented to medical record review were also included in the broader implementation evaluation population. The predominance of PLHIV in the study population reflects the underlying service utilization profile of region F primary healthcare clinics, where HIV-related services constitute a substantial proportion of chronic care visits, rather than sampling intended to achieve balanced representation by HIV status.

In the present secondary analysis, we included both PLHIV and PNLHIV to enable a comparative assessment of progression and regression across stages of the HTN care cascade. Data were extracted for participants enrolled between August 2022 and May 2024, with follow-up through routine medical record review extending to May 2025. Follow-up time therefore varied by enrollment date, with some participants contributing up to approximately three years of observation and others approximately twelve months. This study period allowed assessment of longitudinal transitions across the HTN care cascade within the overall 24-month implementation window and subsequent follow-up period.

### Defining longitudinal HTN care cascade

HTN was defined at baseline by two consecutive, high BP measures or a documented diagnosis of HTN or use of anti-hypertensive treatment from the medical record review at any timepoint. However, in practice, the medical records only documented a single BP reading per visit, and information on whether a second confirmatory measurement was taken was not available. From the BP measures, we were able to *define a category of HTN severity (high, high-normal, normal)*, as follows: normal BP: systolic BP (SBP) 120-129 mmHg or diastolic BP (DBP) 80-85 mmHg; high-normal BP: SBP 130-139 mmHg or DBP 86-89 mmHg; and high BP/HTN: SBP of ≥140 mmHg or DBP of ≥90 mmHg.

*Treated participants* were defined as individuals diagnosed with HTN who were documented in the medical record as taking antihypertensive medication such as hydrochlorothiazide, enalapril, amlodipine, furosemide, or atenolol at any timepoint.

*Diagnosed people* with controlled BP” were defined as individuals diagnosed with HTN and on antihypertensive treatment who had a controlled BP at any timepoint.

*HTN control* was defined as a measured SBP of less than 140 mmHg and a measured DBP of less than 90 mmHg among those who were diagnosed as having HTN [16].

Figure 1 displays the outcomes of the HTN care cascade. We described and compared individuals’ transition through the HTN care cascade during the study period by reporting the progression and regression of HTN care across all clinical transitions:

1. **Undiagnosed:** Defined as a baseline status among participants with high BP at the initial/enrolment visit. Individuals were classified as undiagnosed if they had high BP at baseline and no prior documentation of an HTN diagnosis. The denominator includes all participants with high BP at baseline.
2. **Diagnosed but untreated:** Defined as a baseline status among participants with a documented HTN diagnosis but no evidence of antihypertensive treatment initiation at baseline. The denominator includes all participants diagnosed with HTN at baseline.
3. **Diagnosed and treated:** Defined as participants with a documented HTN diagnosis who had initiated antihypertensive therapy at baseline. The denominator includes all participants diagnosed with HTN at baseline. This definition includes “ever treated” at baseline.
4. **Treated and controlled:** Defined as a longitudinal progression outcome among participants who were on antihypertensive treatment at baseline and achieved BP control at follow-up. The denominator includes all participants treated at baseline, regardless of subsequent regression.
5. **Regressed to untreated:** Defined as a longitudinal regression outcome among participants who were treated at baseline but were no longer on treatment at follow-up. The denominator includes all participants treated at baseline.
6. **Regressed to uncontrolled:** Defined as a longitudinal regression outcome among participants who were treated and controlled at baseline but had uncontrolled BP at follow-up. The denominator includes all participants who were treated and controlled at baseline.

**Figure 1.**
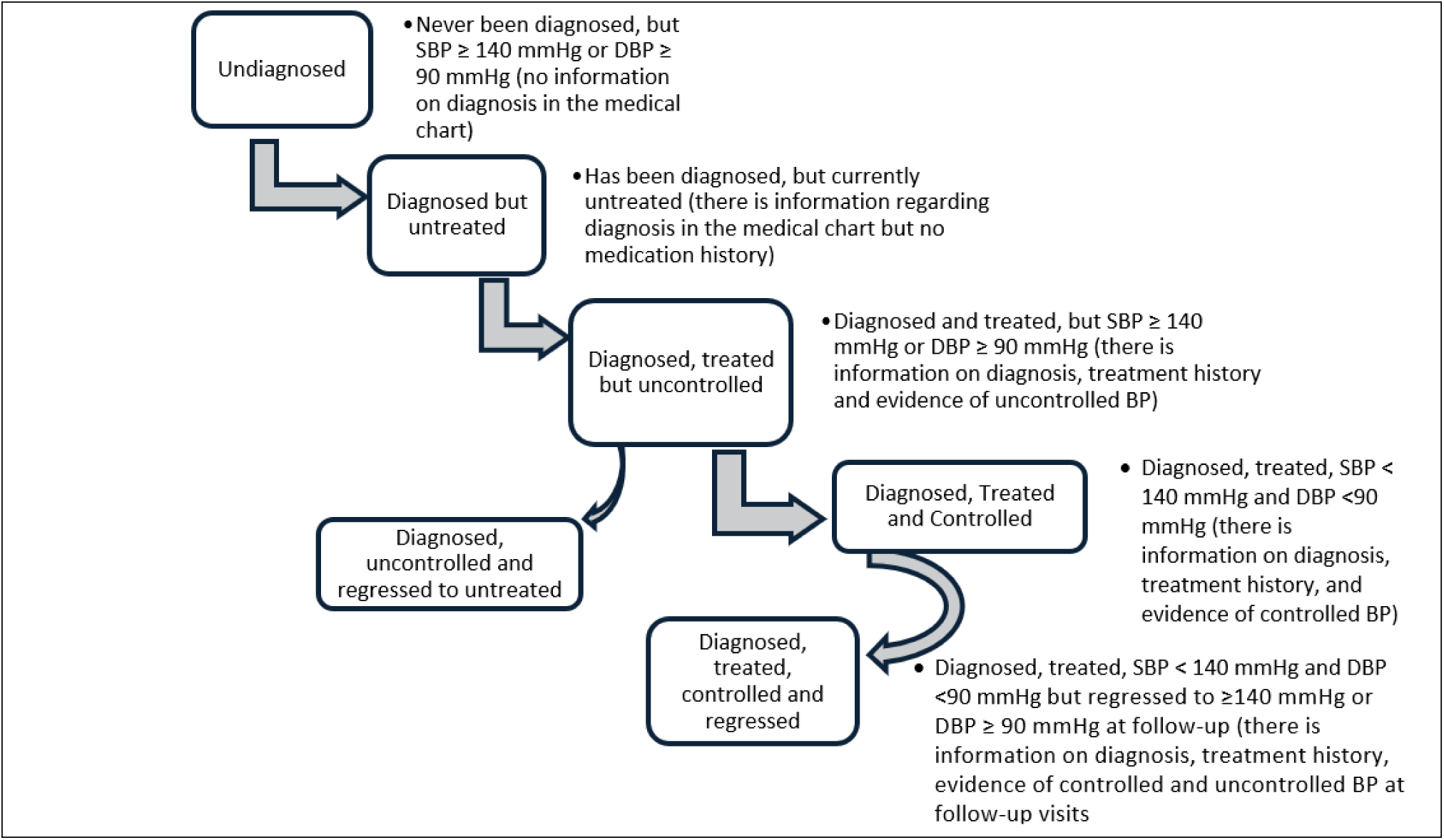
Stages of the HTN care cascade for the study population

### Outcome measurements

#### Primary outcome

Estimates were reported for each outcome (undiagnosed, diagnosed but untreated, diagnosed and treated, treated and controlled, regressed to untreated, regressed to uncontrolled) based on the primary exposure variable (HIV status) and covariates of interest.

“Progression” was defined as movement of individuals from their baseline stage to a more advanced stage of the HTN care cascade at any subsequent timepoint during follow-up. For example, a participant could progress from diagnosis to treatment initiation, or from treatment to BP control. Once BP control was achieved, no further forward progression was possible along the continuum [12].

“Regression” was defined as movement from a more advanced to a prior stage of the HTN care cascade over time, such as loss of BP control or discontinuation of treatment. Because participants were assessed at multiple timepoints, individuals could experience both progression and regression during follow-up. Each change was evaluated sequentially according to the direction of movement between consecutive stages. Once a participant was diagnosed with HTN, they could no longer be considered undiagnosed [12].

### Sample size, inclusion, and exclusion criteria

S1 Fig. presents the flowchart of all participants who met the study’s inclusion criteria (those who consented to participate in the study and whose files were reviewed according to the iHEART-SA trial) [15].

Between August 2022 and May 2024, 37,620 individuals were screened for iHEART-SA trial, of whom 33,484/37,620 (89.0%) had known HIV status. After applying eligibility criteria and obtaining consent, 23,855/25,120 (95.0%) participants ultimately had a medical file or chart review form completed.

### Sociodemographic covariates

The following data were available on sociodemographic covariates: age (categorized as: 18-39, 40-49, 50-59, 60+ in the regression model), sex (male or female), nationality (South African, Zimbabwean, Other).

### Statistical analyses

We reported simple descriptive statistics, frequencies, proportions, and/or medians with interquartile ranges (IQR) for the covariates included in the analyses (sex, age, nationality and BP category). For the tests of associations between HIV status and covariates, the Pearson chi-square test was used in case of dichotomous and categorical variables, and a t-test in case of continuous covariables. We reported the frequencies, and proportions of individuals in the data that made each of the transitions. To calculate the risk magnitude of differences and adjust for different exposures (age, sex, and nationality) or distributions across the longitudinal HTN care cascade stages, we used Poisson regression models for progression and regression through the care cascade with each sociodemographic characteristic as the main exposure. Adjusted risk ratios (aRR) and their corresponding 95% confidence intervals (95%CI) were reported.

## Results

### Sample characteristics

Table 1 provides descriptive characteristics for the study participants. Of the 23,855 participants, 78.5% (18,720/23,855) were PLHIV. The median follow-up duration was 337 days (IQR: 168–532) overall, 336 days (IQR: 168–525) among PLHIV, and 358 days (IQR: 169–559.5) among PNLHIV.

**Table 1:** Descriptive characteristics for the study participants.

| Participant characteristics | All | PLHIV | PNLHIV |
| --- | --- | --- | --- |
| Participants who had their medical records reviewed | 23,855 | 18,720 | 5,135 |
| Median follow-up duration in days (IQR) | 337 (168-532) | 336 (168-525) | 358 (169-559.5) |
| Mean follow-up duration in days (SD) | 369 (1.8) | 368 (2.0) | 372 (3.7) |
| Median age (IQR, in years) | 44 (37-52) | 42 (36-49) | 51 (42-61) |
| Age category (in years) |  |  |  |
| 18-39 | 7,225 (31.42%) | 6,383 (35.50%) | 842 (16.78%) |
| 40-49 | 8,587 (37.34%) | 7,268 (40.42%) | 1,319 (26.29%) |
| 50-59 | 4,946 (21.51%) | 3,518 (19.57%) | 1,428 (28.46%) |
| 60+ | 2,240 (9.74%) | 811 (4.61%) | 1,429 (28.48%) |
| Sex |  |  |  |
| Female | 16,296 (68.31%) | 12,940 (69.12%) | 3,356 (65.36%) |
| Male | 7,559 (31.69%) | 5,780 (30.88%) | 1,779 (34.64%) |
| Nationality |  |  |  |
| South African | 16,097 (67.48%) | 12,380 (66.13%) | 3,717 (72.39%) |
| Zimbabwean | 5,825 (24.42%) | 5,023 (26.83%) | 802 (15.62%) |
| Other | 1,933 (8.10%) | 1,317 (7.04%) | 616 (12.00%) |
| Median SBP (mmHg) for those who underwent file review | 132 (120-145) | 129 (117-141) | 140 (129-154) |
| Median DBP (mmHg) for those who underwent file review | 81 (72-90) | 80 (71-89) | 84 (75-93) |
| Blood pressure categorization* |  |  |  |
| Normal | 10,643 (45.10%) | 9,289 (50.17%) | 1,354 (26.63%) |
| High-normal | 4,821 (20.43%) | 3,810 (20.58%) | 1,011 (19.88%) |
| High | 8,136 (34.37%) | 5,416 (29.25%) | 2,720 (53.49%) |

Abbreviations: BP=Blood Pressure; PLHIV=People living with HIV; PNLHIV=People not living with HIV,
IQR: Interquartile Range; HTN=Hypertension.
BP classification\*: Normal BP: SBP 120-130 mmHg or DBP 80-85 mmHg; High-normal BP: SBP 130-
140 mmHg or 85-90 mmHg; High BP/HTN: SBP of >140 mmHg or DBP of >90 mmHg

**Table 2.**
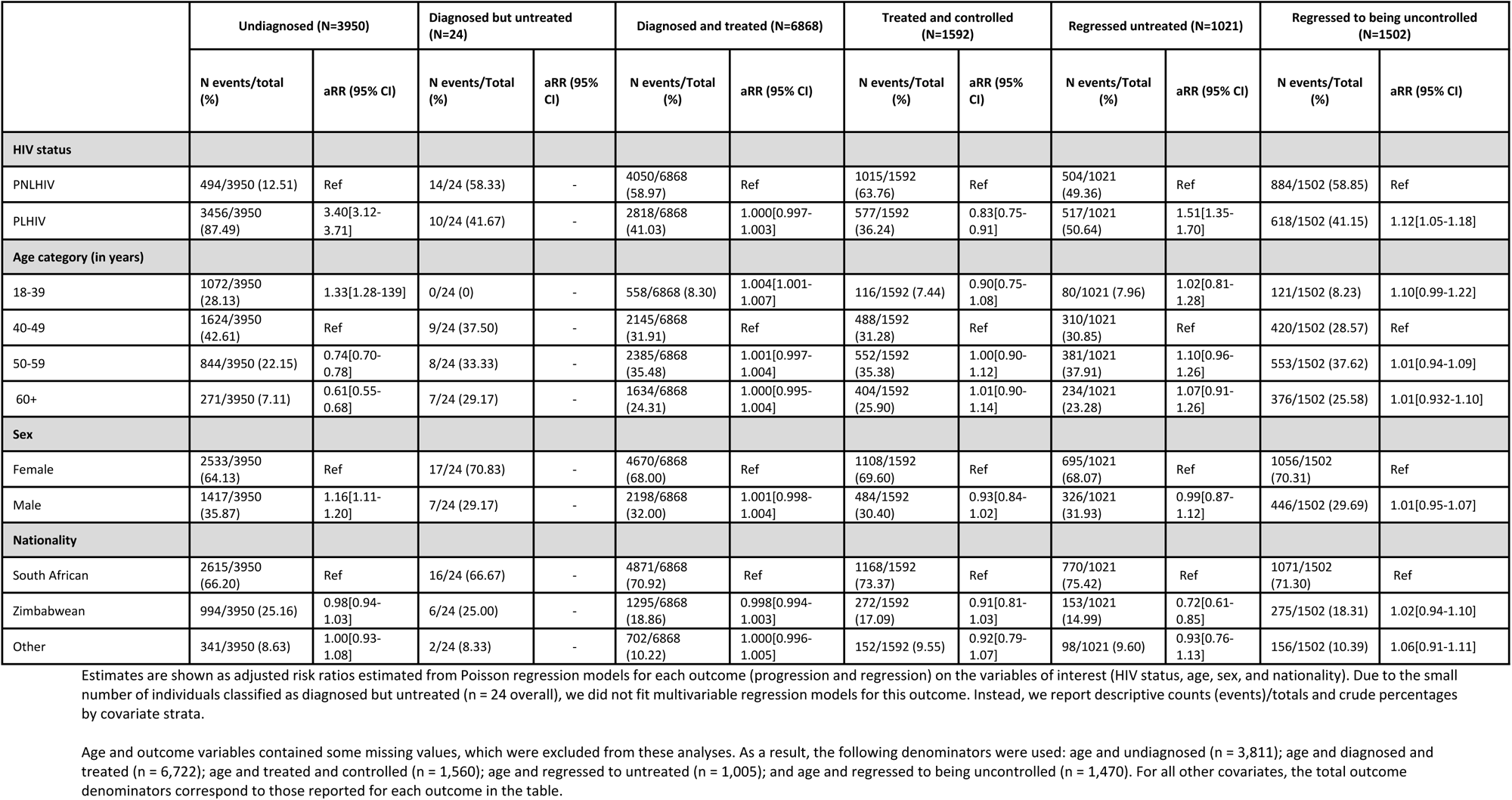
Differences in the adjusted risk ratios (aRR) of progressing and regressing through the HTN care cascade.

|  | Undiagnosed (N=3950) |  | Diagnosed but untreated (N=24) |  | Diagnosed and treated (N=6868) |  | Treated and controlled (N=1592) |  | Regressed untreated (N=1021) |  | Regressed to being uncontrolled (N=1502) |  |
| --- | --- | --- | --- | --- | --- | --- | --- | --- | --- | --- | --- | --- |
|  | N events/total (%) | aRR (95% CI) | N events/Total (%) | aRR (95% CI) | N events/Total (%) | aRR (95% CI) | N events/Total (%) | aRR (95% CI) | N events/Total (%) | aRR (95% CI) | N events/Total (%) | aRR (95% CI) |
| **HIV status** |  |  |  |  |  |  |  |  |  |  |  |  |
| PNLHIV | 494/3950 (12.51) | Ref | 14/24 (58.33) | - | 4050/6868 (58.97) | Ref | 1015/1592 (63.76) | Ref | 504/1021 (49.36) | Ref | 884/1502 (58.85) | Ref |
| PLHIV | 3456/3950 (87.49) | 3.40[3.12-3.71] | 10/24 (41.67) | - | 2818/6868 (41.03) | 1.000[0.997-1.003] | 577/1592 (36.24) | 0.83[0.75-0.91] | 517/1021 (50.64) | 1.51[1.35-1.70] | 618/1502 (41.15) | 1.12[1.05-1.18] |
| **Age category (in years)** |  |  |  |  |  |  |  |  |  |  |  |  |
| 18-39 | 1072/3950 (28.13) | 1.33[1.28-139] | 0/24 (0) | - | 558/6868 (8.30) | 1.004[1.001-1.007] | 116/1592 (7.44) | 0.90[0.75-1.08] | 80/1021 (7.96) | 1.02[0.81-1.28] | 121/1502 (8.23) | 1.10[0.99-1.22] |
| 40-49 | 1624/3950 (42.61) | Ref | 9/24 (37.50) | - | 2145/6868 (31.91) | Ref | 488/1592 (31.28) | Ref | 310/1021 (30.85) | Ref | 420/1502 (28.57) | Ref |
| 50-59 | 844/3950 (22.15) | 0.74[0.70-0.78] | 8/24 (33.33) | - | 2385/6868 (35.48) | 1.001[0.997-1.004] | 552/1592 (35.38) | 1.00[0.90-1.12] | 381/1021 (37.91) | 1.10[0.96-1.26] | 553/1502 (37.62) | 1.01[0.94-1.09] |
| 60+ | 271/3950 (7.11) | 0.61[0.55-0.68] | 7/24 (29.17) | - | 1634/6868 (24.31) | 1.000[0.995-1.004] | 404/1592 (25.90) | 1.01[0.90-1.14] | 234/1021 (23.28) | 1.07[0.91-1.26] | 376/1502 (25.58) | 1.01[0.932-1.10] |
| **Sex** |  |  |  |  |  |  |  |  |  |  |  |  |
| Female | 2533/3950 (64.13) | Ref | 17/24 (70.83) | - | 4670/6868 (68.00) | Ref | 1108/1592 (69.60) | Ref | 695/1021 (68.07) | Ref | 1056/1502 (70.31) | Ref |
| Male | 1417/3950 (35.87) | 1.16[1.11-1.20] | 7/24 (29.17) | - | 2198/6868 (32.00) | 1.001[0.998-1.004] | 484/1592 (30.40) | 0.93[0.84-1.02] | 326/1021 (31.93) | 0.99[0.87-1.12] | 446/1502 (29.69) | 1.01[0.95-1.07] |
| **Nationality** |  |  |  |  |  |  |  |  |  |  |  |  |
| South African | 2615/3950 (66.20) | Ref | 16/24 (66.67) | - | 4871/6868 (70.92) | Ref | 1168/1592 (73.37) | Ref | 770/1021 (75.42) | Ref | 1071/1502 (71.30) | Ref |
| Zimbabwean | 994/3950 (25.16) | 0.98[0.94-1.03] | 6/24 (25.00) | - | 1295/6868 (18.86) | 0.998[0.994-1.003] | 272/1592 (17.09) | 0.91[0.81-1.03] | 153/1021 (14.99) | 0.72[0.61-0.85] | 275/1502 (18.31) | 1.02[0.94-1.10] |
| Other | 341/3950 (8.63) | 1.00[0.93-1.08] | 2/24 (8.33) | - | 702/6868 (10.22) | 1.000[0.996-1.005] | 152/1592 (9.55) | 0.92[0.79-1.07] | 98/1021 (9.60) | 0.93[0.76-1.13] | 156/1502 (10.39) | 1.06[0.91-1.11] |

Estimates are shown as adjusted risk ratios estimated from Poisson regression models for each outcome (progression and regression) on the variables of interest (HIV status, age, sex, and nationality). Due to the small number of individuals classified as diagnosed but untreated (n = 24 overall), we did not fit multivariable regression models for this outcome. Instead, we report descriptive counts (events)/totals and crude percentages by covariate strata.
Age and outcome variables contained some missing values, which were excluded from these analyses. As a result, the following denominators were used: age and undiagnosed (n = 3,811); age and diagnosed and treated (n = 6,722); age and treated and controlled (n = 1,560); age and regressed to untreated (n = 1,005); and age and regressed to being uncontrolled (n = 1,470). For all other covariates, the total outcome denominators correspond to those reported for each outcome in the table.

The median age was 44 years (IQR: 37–52), with PLHIV being younger (median 42 years, IQR: 36–49) compared to PNLHIV (median 51 years, IQR: 42–61). Of the total participants, 68.3% (16,296/23,855) were female, with a slightly higher proportion among PLHIV (69.1%, 12,940/18,720) than PNLHIV (65.4%, 3,356/5,135). South Africans comprised 67.5% (16,097/23,855) participants, more among PNLHIV (72.4%, 3,717/5,135) compared to PLHIV (66.1%, 12,380/18,720). One-quarter of the participants were Zimbabweans (24.4%, 5,825/23,855), with a higher proportion among PLHIV (26.8%, 5,023/18,720) versus PNLHIV (15.6%, 802/5,135).

The median SBP was 132 mmHg (IQR:120-145 mmHg). Overall, 45.1% (10,643/23,600) of participants had normal BP, while 34.4% (8,136/23,600) were classified as having high BP. Among PLHIV, 29.3% (5,416/18,515) had high BP, compared to 53.5% (2,720/5,085) of PNLHIV.

### Movement in the longitudinal HTN care cascade

Results reflect changes in HTN care cascade among participants enrolled in the iHEART-SA trial between August 2022 and May 2024 and followed up through May 2025 using routine medical record data. Progression and regression along the HTN care cascade were therefore assessed for each participant, within the 24-month implementation window and the 12 months follow-up period.

### Progression of individuals across the longitudinal HTN care cascade

Figure 2 illustrates the forward movement of participants through the stages of HTN care. Of the total PLHIV with high BP, 46% (2,998/6,515) were diagnosed with HTN, compared to 91% (3,804/4,200) of PNLHIV (p<0.001). Among those diagnosed, 84% (2,526/2,998) of PLHIV progressed to anti-HTN treatment, compared to 89% of PNLHIV (3,381/3,804) (p=0.137). Among individuals who began treatment, 45% (1,125/2,526) of PLHIV achieved BP control, compared to 63% (2,141/3,381) of PNLHIV, highlighting a significant drop-off at the control stage for both groups, but substantially worse for PLHIV (p<0.001).

**Figure 2:**
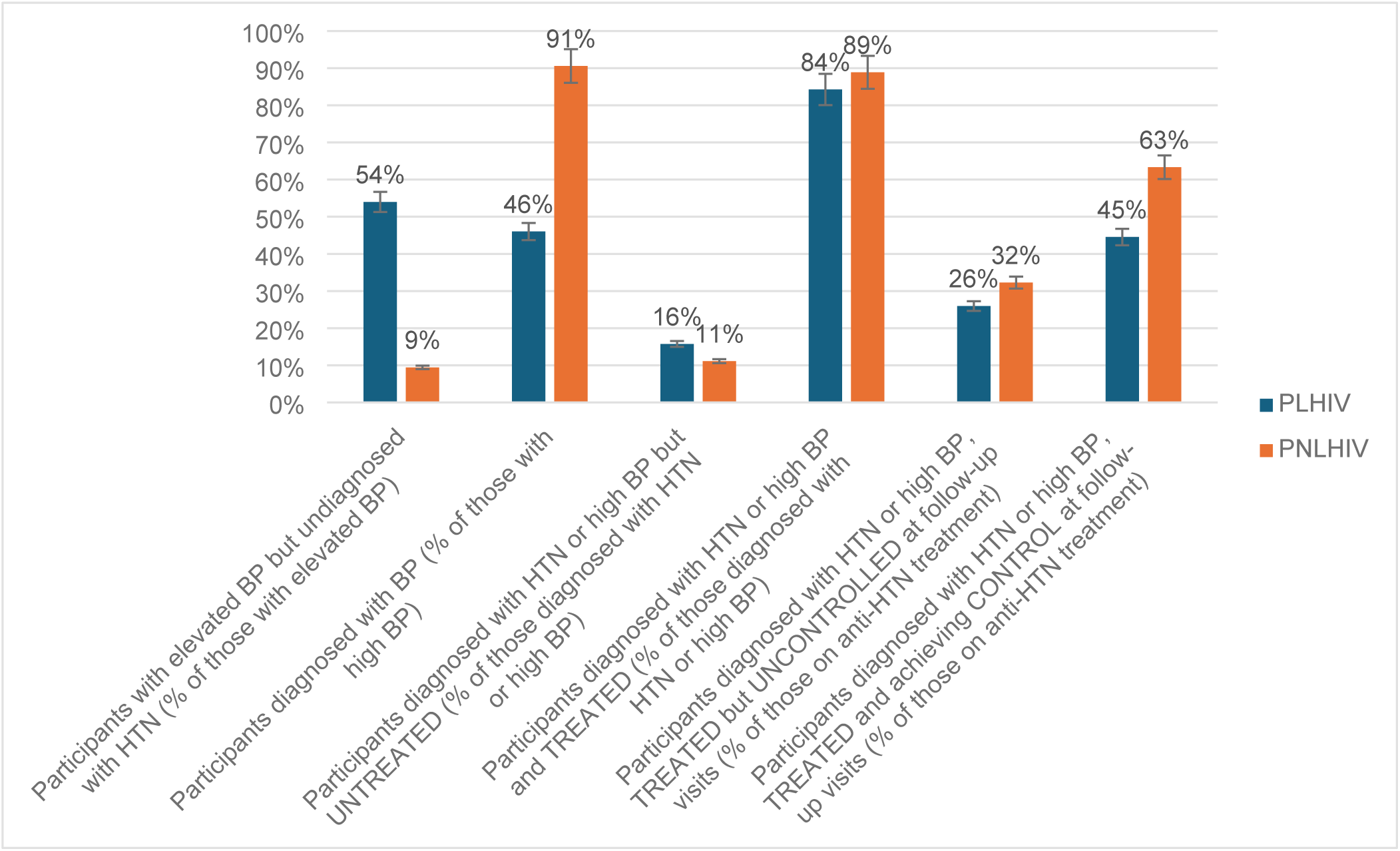
Progression in HTN care cascade among participants enrolled Aug 2022–May 2024 and followed through May 2025

### Regression of individuals across the longitudinal HTN care cascade

Figure 3 presents the regression or backward movement of participants across the cascade. Of those who had been previously diagnosed with HTN and had initiated antihypertensive treatment but remained uncontrolled, almost one in four discontinued treatment and regressed to being untreated at follow-up visits (26%, 392/1524 for PLHIV vs 27%, 522/1922 for PNLHIV) (p=0.469).

**Figure 3:**
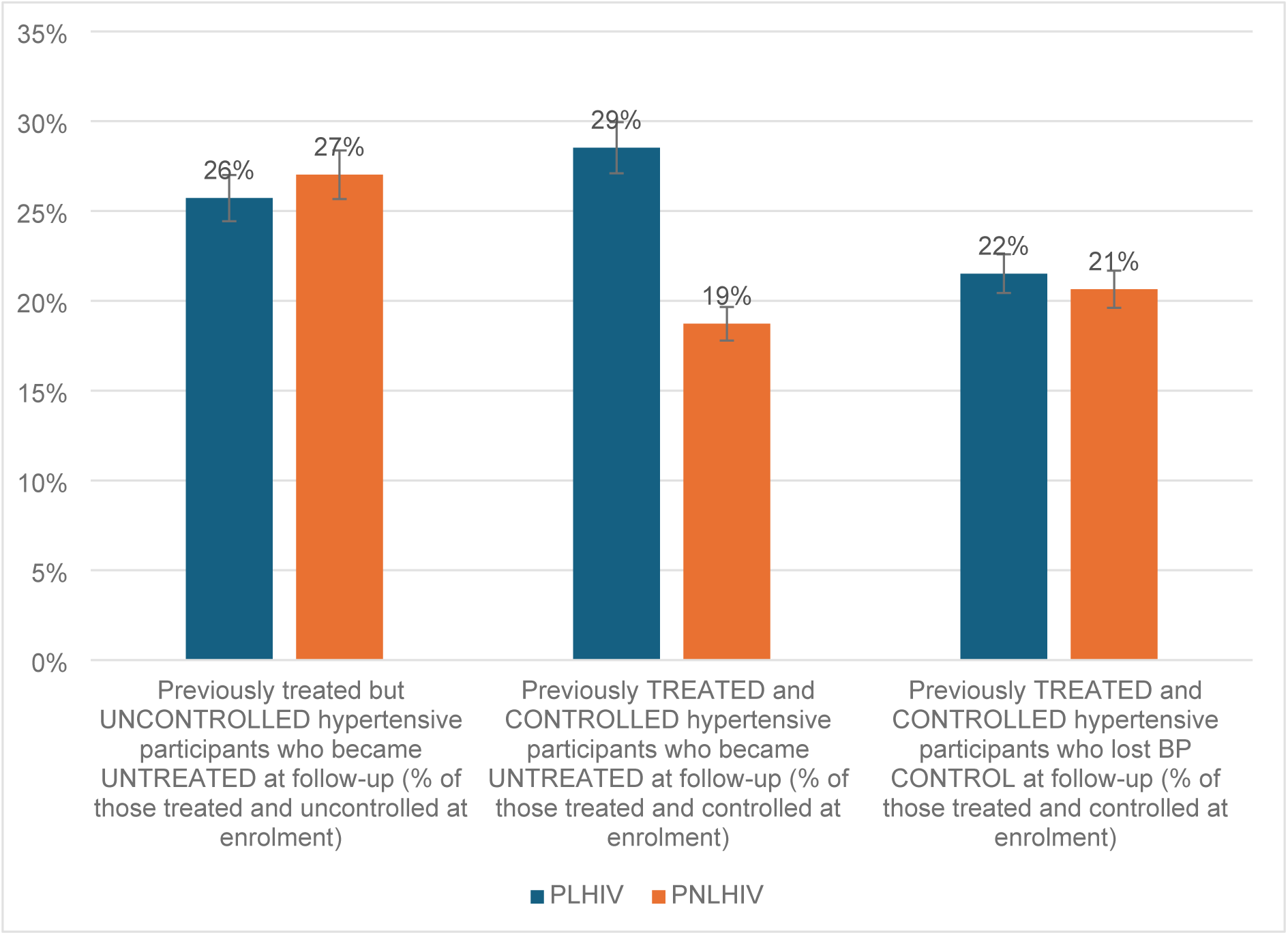
Regression in HTN care cascade among participants enrolled Aug 2022–May 2024 and followed through May 2025

Of those who initially achieved BP control, 29% (243/1125) of PLHIV vs 19% (235/2141) of PNLHIV, regressed to being untreated at follow-up visits (p<0.001). Furthermore, almost one in five of the participants who had achieved BP control were no longer controlled at follow-up (22%, 242/1125 for PLHIV vs 21%, 442/2141 for PNLHIV) (p=0.640).

### Regression analysis Diagnosis of HTN

PLHIV were more likely to remain undiagnosed compared with PNLHIV (aRR 3.40; 95% CI 3.12–3.71). Individuals aged 18–39 years had a higher risk of being undiagnosed with HTN (aRR 1.33; 95% CI 1.28–1.39), while older adults were less likely to remain undiagnosed (50–59 years: aRR 0.74; 95% CI 0.70–0.78; ≥60 years: aRR 0.61; 95% CI 0.55–0.68). Male sex was also associated with undiagnosed HTN (aRR 1.16; 95% CI 1.11–1.20).

### Diagnosed and treated for HTN

Compared with adults aged 40–49 years, participants aged 18–39 years were slightly more likely to be diagnosed and treated for HTN (aRR 1.004; 95% CI 1.001–1.007).

### HTN control among treated individuals

Among those receiving antihypertensive treatment, PLHIV were less likely to achieve BP control (aRR 0.83; 95% CI 0.75–0.91).

### Regression from a more advanced to a prior stage of the HTN care cascade

PLHIV had a higher risk of regressing to untreated HTN (aRR 1.51; 95% CI 1.35–1.70) and of regressing from controlled to uncontrolled HTN (aRR 1.12; 95% CI 1.05–1.18). Zimbabwean nationality was associated with a lower risk of regression to untreated HTN compared with South African nationality (aRR 0.72; 95% CI 0.61–0.85).

## Discussion

This study provides longitudinal evidence on progression and regression across the HTN care cascade among adults with and without HIV in Johannesburg, South Africa. The findings reveal persistent gaps and distinct differences between PLHIV and PNLHIV. HTN prevalence was markedly higher among PNLHIV than among PLHIV, yet rates of HTN diagnosis were substantially lower among PLHIV despite their frequent contact with healthcare services. Although treatment initiation rates were comparable across groups, sustained BP control remained low overall. Follow-up duration did not differ between PLHIV and PNLHIV, indicating that the observed differences in HTN care cascade outcomes are unlikely to be attributable to differential observation time. A higher proportion of PLHIV regressed to being untreated or lost BP control over time, underscoring significant challenges in maintaining continuity of HTN care within HIV-focused health programmes.

The disproportionately higher prevalence of HTN among PNLHIV (53.5%) compared with PLHIV (29.3%) is likely attributable to both demographic and structural factors. PNLHIV in this cohort were considerably older, and age is a well-established determinant of high BP [17]. However, the predominance of PLHIV reflects the service profile of region F primary healthcare clinics, where the majority of chronic care visits are for HIV-related services rather than other chronic conditions. As parent iHEART-SA trial eligibility required participants to be receiving chronic care and to have an existing medical record at the clinic, individuals accessing HIV care were more likely to meet the inclusion criteria and be enrolled. Consequently, PNLHIV may represent a subgroup with a higher burden of cardiometabolic risk, while PLHIV who attend ART clinics might not receive routine HTN screening. This aligns with prior South African evidence demonstrating inconsistent HTN screening among PLHIV despite regular clinic attendance [18].

Our longitudinal results correspond with findings from Mauer et al. [12], who demonstrated substantial treatment discontinuation and loss of BP control across several middle-income countries, underscoring the global challenge of maintaining longitudinal HTN control. In this study, nearly one-third of PLHIV who achieved BP control reverted to being untreated, compared with one-fifth among PNLHIV. This pattern suggests that the most vulnerable point in the HTN care cascade for PLHIV is treatment continuity rather than initiation. Factors contributing to this may include competing clinical priorities within HIV care, therapeutic inertia among healthcare providers, and patient-level barriers such as pill burden and prioritisation of ART adherence [19–23].

Conversely, PNLHIV, who typically access care primarily for HTN, may benefit from more focused and consistent follow-up, leading to better continuity [19,20]. These observations reflect the consequences of health system organisation: vertically oriented HIV programmes achieve exceptional ART continuity but may inadequately support comorbid NCD management. Comparable challenges have been reported in other sub-Saharan African settings, where fragmented care structures have constrained the delivery of integrated chronic disease services [21,22].

In adjusted analyses, HIV status was the most consistent determinant of engagement across the longitudinal HTN care cascade, with PLHIV substantially more likely to remain undiagnosed, less likely to achieve BP control once treated, and more likely to regress to untreated or uncontrolled HTN. These findings suggest that frequent contact with HIV services does not translate into effective HTN detection or sustained management, underscoring persistent gaps in HIV/NCD integration and the need for routine BP screening and longitudinal HTN care within HIV programmes [24–26].

Younger adults and men were more likely to remain undiagnosed, reflecting patterns reported in population-based studies of HTN awareness and diagnosis, and highlighting the limitations of facility-based screening for these groups [27–31]. Although younger adults showed a marginally higher likelihood of being treated, the magnitude of this association was minimal.

Regression after treatment initiation was common, particularly among PLHIV, consistent with prior evidence showing that treatment initiation alone does not ensure sustained BP control in routine care settings, especially in the absence of integrated services and long-term adherence support [30,32–34].

The lower risk of regression to untreated HTN observed among Zimbabwean participants may reflect greater continuity of care and sustained engagement once treatment is initiated, patterns that have been observed in other studies of chronic disease care among migrant populations in Southern Africa [35,36]. Similar findings in NCD cascade analyses suggest that, despite barriers to initial access, non-national populations may experience comparable or improved retention once engaged in care; however, the mechanisms underlying these differences warrant further investigation.

The overall implications of these findings extend to ongoing debates regarding the optimal model of HTN service delivery in high HIV-burden settings. Integration of HTN management into HIV care could leverage existing infrastructure, workforce, and adherence support systems, enhancing efficiency and patient convenience [18,37–39]. Evidence from other integrated models demonstrates improved screening and follow-up when NCD services are embedded within HIV care [18]. However, given the scale and complexity of the HTN epidemic, vertical or semi-vertical programmes with dedicated resources and supply chains may offer advantages in focus and sustainability. The recent home-based HTN care trial in rural South Africa [40], illustrates the potential of decentralised, patient-centred approaches to improve BP control independent of HIV care platforms. Together, these models suggest that effective HTN control will likely require flexible, context-specific strategies that blend integration with differentiated service delivery.

Our study had several limitations. Data were collected from routine medical records, which may be subject to poor recording, and incomplete documentation. For example, BP measurements and HTN diagnoses and treatment initiation may not always be recorded accurately or completely, and or discontinuation of treatment could be under- or over-reported.

Our study did not account for several potentially important determinants of HTN care, including medication adherence, patient socioeconomic status, provider-level factors such as training or attitudes toward integrated care, and clinical covariates such as BMI, which were not collected as part of the parent study. The absence of this data limits the ability to fully describe observed differences in progression and regression between PLHIV and PNLHIV. Nonetheless, some of these determinants or factors have been explored and reported in a qualitative study conducted in this setting [38].

Another limitation of this analysis is the variation in follow-up duration among participants. Enrollment spanned from August 2022 to May 2024, and follow-up extended through May 2025, meaning some participants contributed up to three years of observation while others had about 12 months. This unequal exposure period may have influenced observed rates of progression or regression along the HTN care cascade, as earlier enrollees had greater opportunity to initiate or achieve BP control. However, despite this variability, there was no statistically significant difference in follow-up duration between PLHIV and PNLHIV, suggesting that differential exposure time is unlikely to have biased comparisons between the two groups. The observed patterns remain informative of real-world dynamics in care engagement across time and highlight how longitudinal retention shapes outcomes in routine program settings.

Our analysis was also limited by the challenge of capturing movement across the HTN care cascade. Participants could experience both progression and regression during follow-up, and the timing between observations varied. As a result, transitions between cascade stages may not fully reflect the true continuity or duration of engagement in care. Consequently, our estimates of movement along the cascade should be interpreted as indicative of stage changes between recorded visits rather than continuous trajectories over time.

The study sample was drawn from a single urban setting in South Africa, which may restrict the generalizability of results to other populations or regions with different healthcare infrastructure or patient demographics. Given that study participation and follow-up depended on engagement with the healthcare system, individuals who are less likely to seek care or who disengage from care may be underrepresented. This could lead to an overestimation of progression rates and an underestimation of regression.

In addition, the overrepresentation of PLHIV reflects the underlying service utilisation profile of region F primary healthcare clinics, where the majority of chronic care visits are for HIV-related services rather than other chronic conditions. This sampling bias, arising from clinic-based recruitment within chronic care services, should be considered when interpreting comparative findings between PLHIV and PNLHIV.

As with many studies using routine data, there is the possibility of missing data or measurement errors in BP readings, diagnosis, diagnosis dates, or treatment status. This might contribute to bias in our results. While this study offers valuable longitudinal insights into the longitudinal HTN care cascade for PLHIV and PNLHIV, its findings should be interpreted in light of these methodological and contextual limitations.

## Conclusion

This study identifies substantial gaps across the longitudinal HTN care cascade among adults attending urban primary healthcare clinics in South Africa, with pronounced disparities among PLHIV. Despite frequent contact with health services, less than half of PLHIV with high BP were diagnosed with HTN, and only 45% of those treated achieved BP control. High levels of regression, including treatment discontinuation and loss of control, were observed across both PLHIV and PNLHIV. These findings underscore the need for comprehensive, integrated and targeted chronic care models that strengthen HTN screening, diagnosis, and sustained control within routine HIV services. Future research integrating more comprehensive patient- and provider-level data is needed to better explain drivers of progression and regression within the HTN care cascade.

## Acknowledgements

We would like to acknowledge the iHEART-SA field team for collecting the data. We would also like to thank the health workers in the 9 PHC facilities in the Johannesburg CBD and all the study participants, without whom this work would not be possible.

## Author contributions

SG, JMG, EKO, NB, AB, SL-E contributed to the study conception and design. SG performed data analysis and interpretation of data for the work. SG wrote the manuscript draft. All authors [SG, JMG, EKO, NB, AB, SL-E] critically commented on the article and read and approved the final manuscript.

## Data availability

Data is available from the corresponding author on reasonable request.

## Ethical approval

This was a secondary analysis of data from the iHEART-SA trial which received approval from the Human Research Ethics Committee (Medical) of the University of Witwatersrand (M211160) and the Johannesburg District Research Office (NHRD No. GP_202111_015). This study was also approved by the Human Research Ethics Committee (Medical) of the University of Witwatersrand (M2411128).

